# Resilience of health systems in Africa to infectious disease shocks: A systematic review

**DOI:** 10.1101/2025.10.03.25337229

**Authors:** Denis Okethwangu, Marit Johansen, Sherry Ahirirwe, Mahima Venkateswaran, Roy Mayega, Elizeus Rutebemberwa, Felix Ocom, Victoria Nankabirwa, Charles L. Okot, Evelyne B. Nyachwo, Sandra Nabatanzi, Evelyn Asio, Umaer N. Mohammed, Alex R. Ario, Frode Forland, Suzanne N. Kiwanuka

## Abstract

Stronger health systems are better equipped to withstand shocks and continue providing quality services as response measures are implemented. We conducted a systematic review to synthesize the understanding of the concept of health system resilience from various stakeholders in Africa, focusing on definitions and attributes of a resilient health system. We conducted a search for peer-reviewed articles and grey literature, filtered for Africa, from 1980 to 2023, using the SPIDER framework. We searched four databases: PubMed, the Bielefeld Academic Search Engine, the Cumulative Index to Nursing and Allied Health Literature, and Scopus, and reviewed the websites of the World Health Organization, Africa CDC, and Ministries of Health of African countries. Articles were selected based on set inclusion and exclusion criteria. Qualitative articles were appraised using the Critical Appraisal Skills Programme, and mixed-methods articles using the Mixed Methods Appraisal Tool. We mapped the distribution of included articles by country studied; categorized the articles based on reported shock, health system building block described; and identified the definition of health system resilience, and its attributes in each article. The search yielded 4,306 relevant records, fifty-five of which were included in the study. Studies were found from 48 of the 54 African countries. Up to 75% of the articles focused on COVID-19; others were on Ebola Virus Disease, cholera, and meningitis. Service delivery and health workforce were the most frequently studied health system building blocks. In defining or describing health system resilience, the adaptive capacity (39, 65%) was most frequently mentioned, followed by absorptive capacity (17, 28%), preparedness (3, 5%), and recovery (1, 2%). Identified attributes of a resilient health system were: community engagement and involvement; leadership and governance; collaborations and partnerships; human resources for health; health education and promotion; health information systems; health service delivery; decentralization and local governance; health infrastructure and logistics; preparedness; learning and adaptation; and innovation and financing. Our review reports four core capacities that define a resilient health system: preparedness, absorptive capacity, adaptive capacity, and recovery. Essential attributes encompass community engagement, health education and promotion, leadership and governance, surveillance and laboratory capacity, innovation, service delivery, and adaptability.

## Introduction

The normal functioning of health systems is disrupted during significant shocks, primarily owing to the pressures these shocks place on various components of the system (1). Documented shocks encompass infectious disease outbreaks, large-scale population displacements, civil unrest, and issues related to climate change (2–4). These shocks diminish the system’s ability to provide high-quality routine healthcare and to respond to emergencies effectively. Disruptions within health facilities frequently arise from the reallocation of resources, whether human, financial, or material, to crisis response efforts, thereby diverting resources from service delivery, logistics, or influencing community perceptions (3). On a larger scale, systemic issues, including governance deficiencies, insufficient financing, structural and resource limitations, and fragile information infrastructure, further intensify disruptions (5). Such disruption may result in an indirect loss of lives, economic downturns, social upheavals, and diminished access to quality services in specialized clinics (e.g., non-communicable diseases, reproductive health, and HIV), which are frequently deprioritized during outbreaks (6,7). In developed economies, the COVID-19 pandemic exposed the insufficiency of access to universal health coverage, which is comprehensively defined as: 1) an adequate number of trained health professionals; 2) availability of medicines; 3) resilient health information systems, including surveillance mechanisms; 4) suitable infrastructure; 5) adequate public financing; and 6) a robust public sector capable of providing equitable and high-quality services (8,9). In developing countries, especially in African nations within the meningitis belt and regions endemic with arboviruses, multiple outbreaks are frequently observed (10,11). Since 2020, the African region has faced more than 100 major outbreaks annually, and Uganda alone experienced over 800 between 2000 and 2022 (12,13). Despite recurring crises, anecdotal evidence indicates that health systems in numerous African nations continue to lack sufficient resilience to respond effectively to emergencies while maintaining routine operations and care (14,15). Given the rising frequency and increasing severity of infectious disease outbreaks, it is essential to strengthen health systems to effectively respond to emergencies. Resilience is defined as “the capacity of health actors, institutions, and populations to prepare for and effectively respond to crises; maintain core functions when a crisis hits; and, informed by lessons learned, reorganize if conditions require it” (16). The concept gained prominence during the Ebola Virus Disease outbreak in West Africa, when health systems lost the ability to ensure health for all and achieve good outcomes (17). Similarly, the COVID-19 pandemic revealed extensive challenges in establishing resilient systems (18–20). To establish resilient health systems on a global scale, it is essential to understand the concept of resilience within specific contexts. For Africa, this necessitates analyzing the concept through the perspective of local stakeholders. Accordingly, a systematic review of published literature indexed in prominent databases was conducted. Our primary objective was to examine evidence regarding health system resilience in African nations, with particular aims of synthesizing the various interpretations of resilience and identifying characteristics of resilient systems in response to infectious disease shocks.

## Materials and methods

### Search and screening

Our review was written in accordance with PRISMA Reporting Guidelines (21) (see Supporting Information, *S1 Checklist*).

We developed a step-wise search strategy using keywords in our review strategy, initially in PubMed, which we modified for the other databases.:

Search #1: resilience

Search #2: health system OR healthcare systems

Search #3: pandemics OR disease outbreaks

Search #4: Africa

Search #5: study type

Search #6: health systems response to disease outbreaks in Africa—#2 AND #3 AND #4

Search #7: Qualitative and mixed studies on health systems response to disease outbreaks in Africa—#6 AND #5

Complete search (#8): #7 AND #1

We combined the different searches to derive the complete set of search terms and results. With the keywords, PubMed generated additional related terms using the exploded medical subject headings (MeSH) function. Using Boolean operators and truncations, we adopted the search terms from PubMed for the other databases depending on the limits to number of search terms and unique search features. For search #4, we adopted the search filter for the African continent developed by Pienaar et al. (21). The other electronic databases we searched were: the Bielefeld Academic Search Engine (BASE), Medline, the Cumulative Index to Nursing and Allied Health Literature (CINAHL), and Scopus. The search strategy development was guided by the Sample, Phenomenon of interest, Design, Evaluation, and Research type (SPIDER) framework. The SPIDER framework was developed as an alternative to the Population, Intervention, Comparison, and Outcome (PICO) framework to guide search strategies in qualitative and mixed-methods systematic reviews (22).

We searched the databases from November 2023 to February 2024. PubMed, NLM (pubmed.ncbi.nlm.nih.gov/) was searched on November 20, 2023; CINAHL accessed through EBSCO was searched on February 28, 2024; Scopus accessed through Elsevier was searched on February 28, 2024; and BASE (www.base-search.net) was searched on March 20, 2024. We also explored websites of organizations like the World Health Organization (www.who.int/), US Centers for Disease Control and Prevention (US CDC) (www.cdc.gov), and Africa Centers for Disease Control and Prevention (AfCDC) (africacdc.org) for grey literature. Additional articles were found from the reference lists of selected articles. Details of the search strategy are in Supporting Infomration (*S1 Text*).

We imported all search results from every electronic database into Zotero, where we identified and removed duplicates (23). We then exported the selected records into the free web version of Rayyan for title and abstract screening (24). The first author (DO) and the third author (SA) conducted blinded title and abstract screening. Where there was a disagreement about whether to include or exclude a record, a decision was made by consensus. In cases where there was no consensus, a senior team member was consulted. We obtained the full texts of articles that passed the title and abstract screening and uploaded them into Covidence. This web-based collaboration software platform streamlines the production of systematic and other literature reviews (25). Articles where full texts could not be retrieved were excluded from the full-text screening. Authors DO and SA independently reviewed the full texts, including or dropping articles based on the inclusion and exclusion criteria.

### Inclusion and exclusion criteria

Peer-reviewed journals articles reporting primary data on health system resilience to infectious diseases conducted from 1980 were included. The cut-off year was selected in order to include any earlier studies on resilience in the health system, even though the concept took root in the health sector in the early 2000’s. Our inclusion criteria were: 1) Articles on health system resilience to infectious disease shocks; 2) Studies conducted in an African country; 3) Articles written in English; and 4) Articles that are qualitative or mixed methods studies. Articles were included if they met these criteria and were conducted from 1980. We excluded studies that examined the resilience of health systems to conflict, climate change, or shocks other than infectious diseases, as well as those that reviewed other studies. We excluded quantitative studies, opinions, commentaries, and editorials because they lack primary qualitative data to support interpretive synthesis. All excluded records are included in Supporting Information (*S1 Table*).

### Data extraction

Our data extraction was conducted in Covidence (25). The data extraction tool was developed in MS Excel by DO, with technical reviews from MJ, MV, AAR, and SNK. In addition to the bibliometric data (title of the article, author, and year of publication), we also extracted data on: the country or countries where the study was conducted, the definition or description of health system resilience, the characteristics and attributes of a resilient health system, and the shocks studied. Data extraction was also conducted independently DO and SA. A consensus was used to reach an agreement in cases of disagreement during the data extraction process.

### Quality assessment of reports

Articles selected from the full-text screening were assessed for quality using the Critical Appraisal Skills Programme (CASP) checklist for qualitative studies, while for mixed-methods studies, we used the Mixed Methods Appraisal Tool (MMAT) checklist (26,27). These two checklists were uploaded onto the Covidence to facilitate the appraisal process. The CASP checklist evaluates three broad areas systematically through a 10-question format. These issues include: 1) Are the results of the study valid? 2) What are the results? 3) Will the results be applicable locally? Each question offers three response options: “yes”; “can’t tell”; and “no”. Prompts, also known as hints, help guide responses to each question. To quantitatively assess the quality of the articles, we assigned a score of 1 to every “yes” score; 0 to “can’t tell”; and -1 to “no”. This was a modification to a study that assigned 2 to every “yes” response; 1 to “can’t tell”; and 0 to “no” (28). Based on total scores, studies were stratified into three quality tiers: high (7–10), moderate (5–6), and low (<5). This scoring approach, while not prescribed by CASP, aligns with practices described by a study that advocates for pragmatic adaptations of the tool to enhance its usability in qualitative evidence synthesis (29). The MMAT is a critical appraisal tool designed to evaluate, among other things, the quality of mixed methods studies. It can be used to assess the quality of primary studies based on experimental or observational data. When appraising mixed-methods studies, the rationale for using a mixed-methods approach is first examined, followed by separate evaluations of the qualitative and quantitative components. In line with guidance from the MMAT developers, we did not calculate an overall numeric score, in order not to oversimplify nuanced methodological judgments (27). Instead, we reported criterion-level ratings and synthesized findings narratively, highlighting recurring strengths. This approach allowed us to transparently interpret the methodological rigor of each study and consider its influence on the synthesis. Articles that scored between 11 to 15 were categorized as high quality; 7 to 10, moderate; and less than 7 were considered low.

### Data management and analysis

We conducted a reflexive thematic analysis (30). From the extracted data, we generated codes that related to our research questions. These codes were then grouped into categories based on their relationships with each other. For the attributes of a resilient health system, twenty-eight categories emerged from the codes that were generated from the extracted data. Using a thematic framework, these categories were further categorized into themes, from which we derived our attributes of a resilient health system. We used Microsoft Excel to describe the articles by country, study type, building block of the health system studied, and shock studied. We used QGIS to map the countries whose health systems were studied. Data were analyzed using NVivo 15.

## Results

### Description of included studies

The search across various electronic platforms yielded a total of 4,306 records, including 1,636 from grey literature and 222 from forward citations. We removed 1,090 duplicate records and screened 3,216 for relevance based on titles and abstracts. During screening, we excluded 2,852 records for reasons such as inappropriate population, outcome, studies conducted outside Africa, language other than English, unsuitable study design, publication type, and shocks other than infectious diseases. The title and abstract screening resulted in 364 records for full-text review. During full-text screening, 118 records (32%) were excluded because the PDFs were not retrievable. Of the remaining 246 records, 131 were identified as duplicates by Covidence, and three were deemed irrelevant. We ultimately screened 112 records, from which we included 55 for review. The PRISMA flowchart, detailing the process, is shown in Fig 1 below. The included papers are in Supporting Information (*S2 Table*).

**Fig 1.**
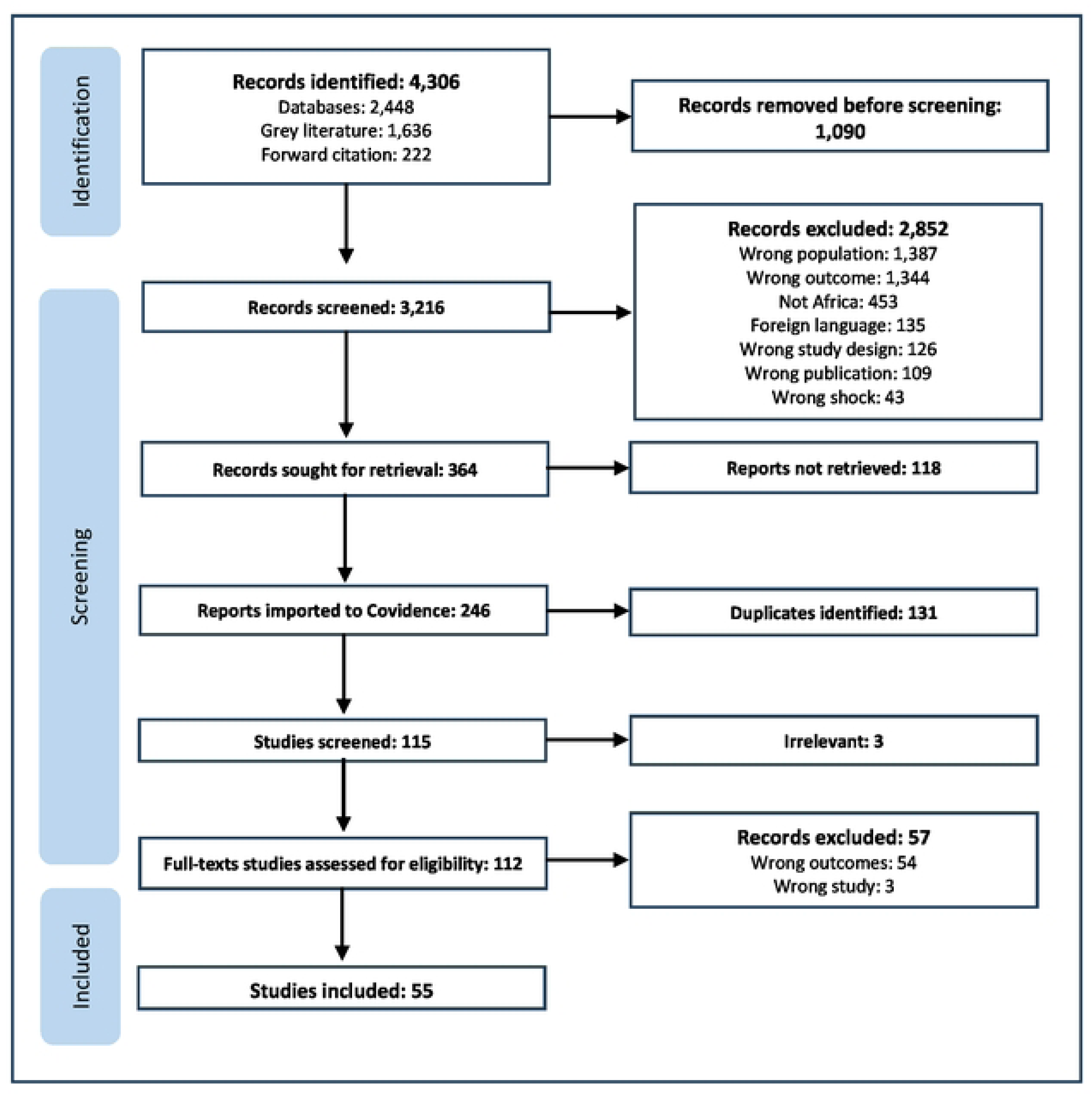
The PRISMA flowchart of studies searched and included.

**Table 1.** Articles included in the systematic review.

Almost three-quarters (74.5%) of the included articles focused on COVID-19 (Table 2). This was followed by Ebola (20%). Of the 54 African countries, 48 were represented in the included articles (Fig 2). Up to 50 of the 55 included articles (91%) focused on service delivery as a health system building block, 45/55 (82%) addressed human resources for health; health information systems was the least discussed building block, featuring in only 17 of the 55 (31%) articles (Fig 3).

**Fig 2.**
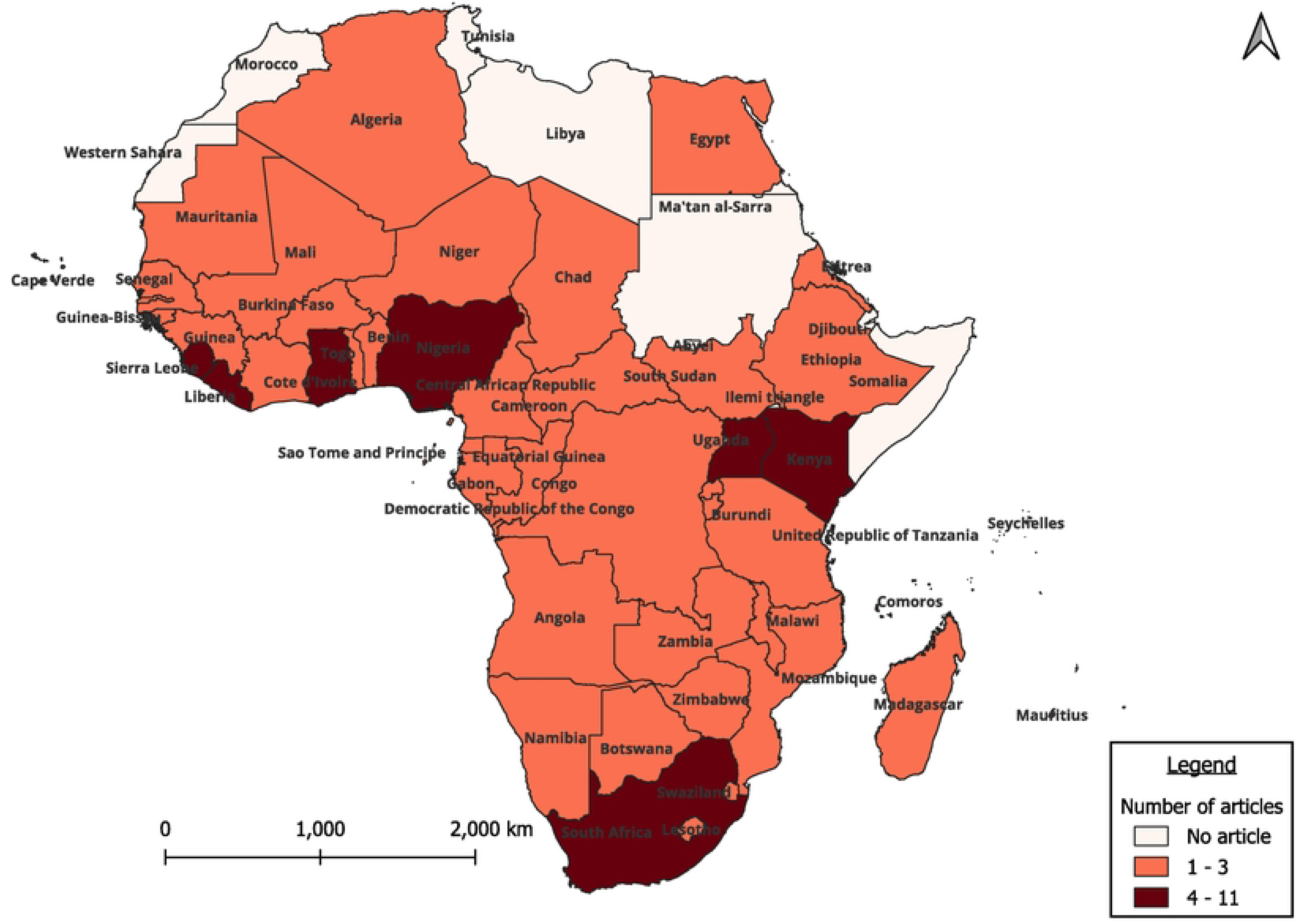
Countries whose health systems were studied in included papers.

**Fig 3.**
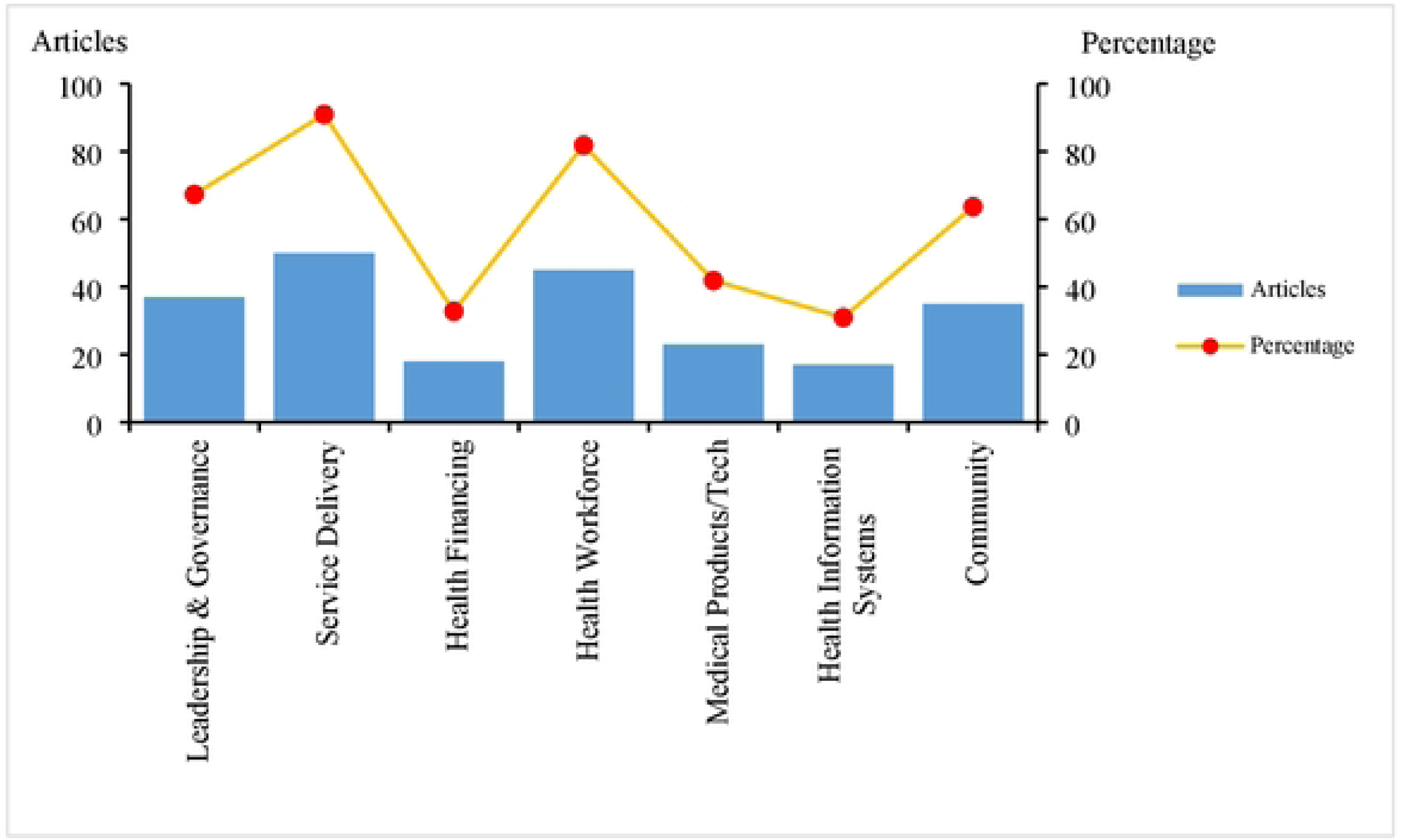
Health system building blocks studied in selected articles.

**Table 2.** Study types among selected articles.

| Study type | COVID-19 | Ebola | Infectious diseases (other) | Cholera | Total |
| --- | --- | --- | --- | --- | --- |
| Mixed methods | 13 | 5 | 1 | 1 | 20 |
| Qualitative | 28 | 6 | 1 |  | 36 |
| Total | 41 | 11 | 2 | 1 | 55 |

### Categories generated from themes

Out of 55 papers included, 46 (83.6%) discussed the description or definition of health system resilience. A critical review of the articles identified four main themes in how resilience is described or defined. These themes include: preparedness, absorptive capacity, adaptive capacity, and recovery. We identified seven attributes of a resilient health system from the themes identified in the data extracted from the included articles (Table 3).

**Table 3.**
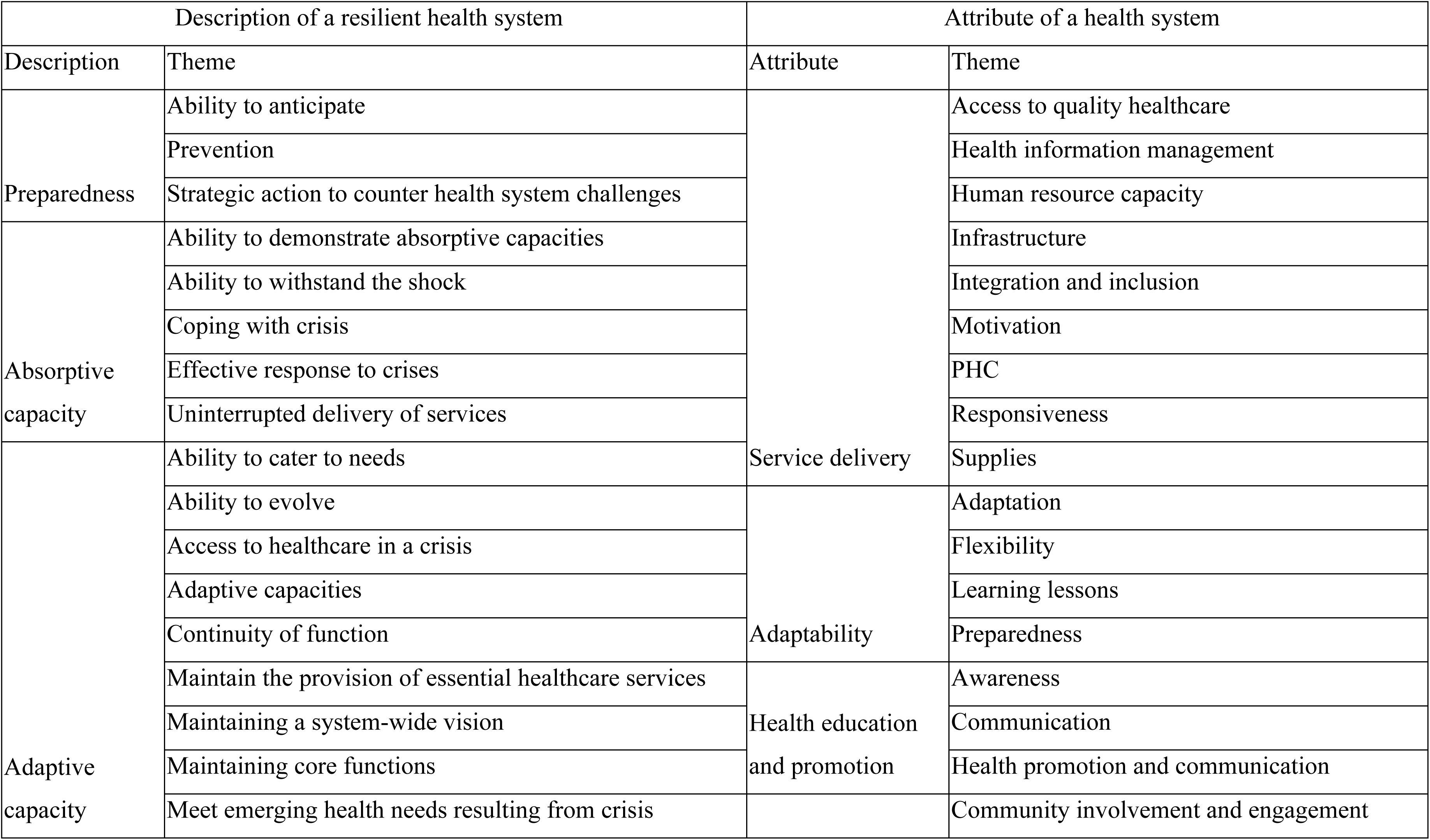
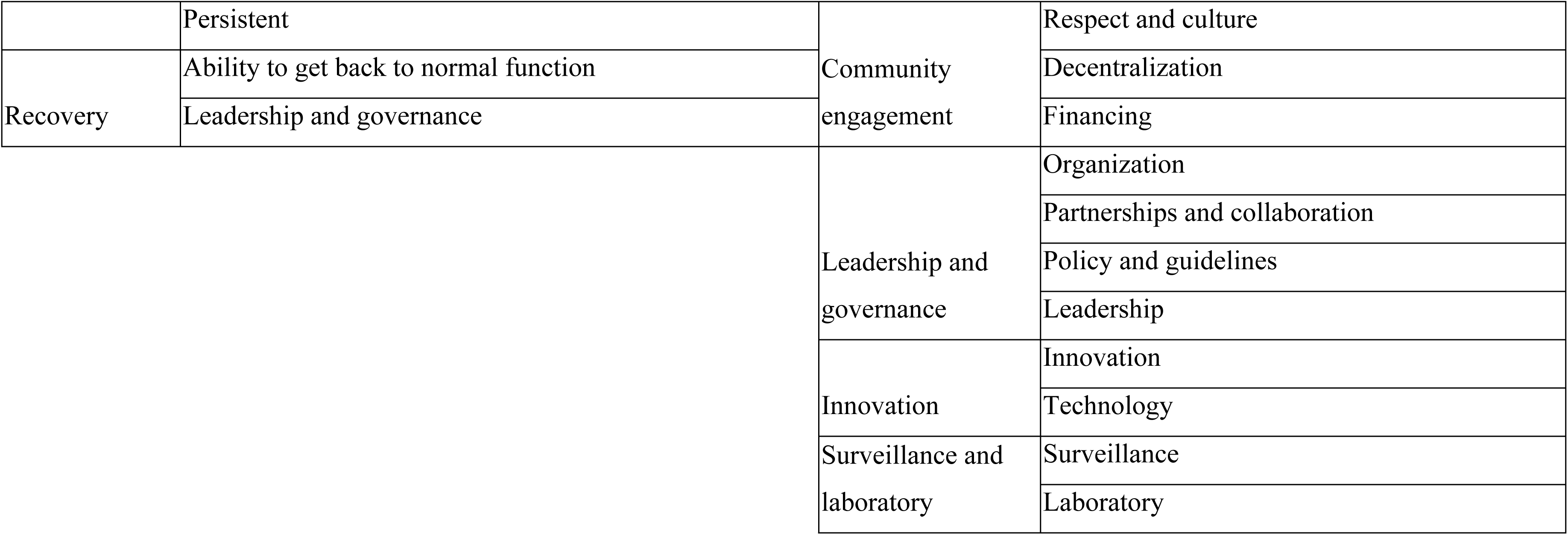
Categories generated from themes.

### Definition of health system resilience

Below, we describe the main categories that emerged from the extracted data and may be considered the capacities that define health system resilience to infectious disease shocks in Africa.

#### Preparedness

The intentional precautionary measures taken to strengthen a health system in anticipation of imminent public health threats are what constitute preparedness (31). Health systems that exhibit great preparedness capacities also put in place prevention measures as a way of averting crises (82).

#### Absorptive capacity

The ability of a health system to continue providing essential health care services while responding to a shock highlights its absorptive capacity. The resilience of a health system refers to its capacity to maintain continuous healthcare services during and after a crisis. (78). This capacity for effective response reflects the health system’s ability to withstand shocks or manage a crisis it encounters (34,53,84).

#### Adaptive capacity

The adaptive capacity of a health system includes its ability to anticipate challenges and put in place appropriate mechanisms to respond to them effectively (43,64). Health systems, while they respond to emergencies, should have the ability to provide basic, as well as emerging, health needs during the response period (83). They should be able to maintain their core functions, as well as hold on to a system-wide vision during a crisis (31,59,76). Where physical access is not possible, as in lockdowns, there should be innovative ways of ensuring that essential healthcare services are available to all who need them (80). Health systems should be able to evolve from their pre-crisis state to develop structures and systems that can withstand future events or put in place innovative measures to ensure that they fight back against the crisis while at the same time maintaining their core functions (61,79).

#### Recovery

Health systems that can bounce back after a crisis and continue providing a similar quality of health services during, as before a crisis (40,64,79,84).

### Attributes of resilience

Below are the categories that we considered the attributes of a health system resilient to infectious disease shocks.

#### Community engagement and involvement

Community involvement in service delivery empowers residents to protect their own health (84). Communities can participate in designing primary healthcare programs, raising awareness about prevention, and assessing resilience initiatives (50,51,56). During emergencies, they play a key role in risk communication, guiding response teams, contact tracing, and reporting cases (39,45,49). Such participation ensures culturally sensitive responses and fosters trust between health systems and communities (53,63).

#### Leadership and governance

Leadership and governance are essential for policy development, enforcing strategic guidelines, and establishing regulatory mechanisms (20,84). They ensure the availability of protocols and emergency frameworks that guide resilient operations (31,75), and coordinate alignment among stakeholders, including the private sector, international partners, health workers, and communities (31,42,46,49).

In Tanzania, government–private sector cooperation, as a leadership function, strengthened the resilience of pharmaceutical supply chains (84). During outbreaks, such partnerships create multidisciplinary response teams, mobilize resources, and leverage expertise from governments, civil society, and community-based organizations (37,49,51,64,66).

Financing underpins resilience, but health services often face underfunding, especially at the community level where workers and programs remain unsupported (43,52,53,69,80). In Uganda, filling gaps through government and partner support was recommended (75).

Decentralization enhances subnational systems by giving authority over resource distribution, service delivery, and outbreak response (63,68). This bottom-up approach promotes efficient decision-making and close supervision, as demonstrated in Cameroon where decentralized responses improved resilience (41,68).

#### Learning and adaptation

Adaptability is crucial for strengthening a health system’s resilience (20). It allows systems to respond to crises while maintaining essential functions (42,46). Adaptive systems are flexible, capable of balancing external demands with quick internal reorganization (61). For instance, integrating programs promotes interdependence and efficient resource use (42,45,81). Learning from past activities further enhances adaptive and transformative capacity (45,54,77). Preparedness involves deliberate precautionary actions taken before a threat, such as training, developing protocols, and stockpiling measures (46). In Namibia, preparedness and planning helped sustain pharmaceutical and supply chains during COVID-19 (70).

#### Health service delivery

Access to quality healthcare is fundamental to system resilience (40,50,65). Emergencies such as lockdowns and travel bans limit access, but innovative approaches are needed to maintain service continuity (44,75). Primary healthcare (PHC), essential to universal health coverage, requires sufficient funding and workforce investment to uphold resilience (81). Resilient systems stay responsive, timely, integrated, and inclusive (51,82). With regards to health infrastructure— physical facilities, equipment, and essential supplies—COVID-19 highlighted the importance of timely access to protective equipment, diagnostics, and medical technologies (43,52,68,69,75,84). Health information systems enhance coordination and efficiency through effective data sharing (31,36,40,43,45,46). Human resources for health are another cornerstone. Emergencies often divert staff from essential services, requiring innovative mechanisms such as surge rosters, redistribution, or reallocation of staff (51,55,81). Capacity-building enhances preparedness, while teamwork fosters efficiency and solidarity (31,58,68,72,82). Motivation through adequate incentives is equally vital (58,72,77).

#### Surveillance and laboratory

Quality data supports effective surveillance, which is vital for the early detection of public health threats and guiding prompt responses (20,43,49,81). Keeping laboratories functional during both normal periods and emergencies boosts resilience. Robust laboratory networks and field deployments have proven essential in mounting successful responses to Ebola outbreaks (20,35).

#### Innovation

During the Ebola outbreak in Sierra Leone and the COVID-19 pandemic, technology and innovation proved vital for optimizing programs, planning, and data collection (42). In emergencies, both technology-based and non-technology solutions are essential. Telemedicine and e-platforms help reduce facility crowding, maintain access during lockdowns, and support remote supervision (55,75,76). Non-technological innovations include multi-month drug refills, using patient networks for medicine distribution, and organizing community clinics (44,75).

#### Health education and promotion

Risk communication and health promotion enhanced resilience during COVID-19 in Uganda and among healthcare workers in South Africa (40,63,75). Clear communication and strong health education empower communities with knowledge and awareness, enabling informed choices that support prevention (49,56).

### Quality assessment

Twenty-seven of the thirty-six qualitative studies included were of high quality; five were of moderate quality, and four were of low quality. Over 80% of the studies yielded valid results, as evidenced by responses to questions 1 to 6 of the assessment tool. Additionally, more than 86% of the studies reported considering ethical issues, employing sufficiently rigorous data analysis, and providing clear statements of findings. Based on the local application of these findings, over 90% of the articles were deemed relevant. See Supporting Information (S3 Table).

Among the twenty mixed methods articles evaluated using the MMAT, all articles passed the screening questions 1 and 2. Only one was of a high quality; ten (50%) were of moderate quality; while the rest were of low quality. eleven (57.9%) scored 60% or higher (Table 5). While no papers were discarded on the basis of their lack of quality, their weight in synthesis was handled with caution. See Supporting Information (S4 Table)

## Discussion

Our study found that the description of health system resilience to infectious disease outbreaks relates to the level of preparedness, its capacity to absorb an outbreak when it occurs, its ability to adapt and sustain quality healthcare services during a response, and its capacity to recover after the crisis. We interpret these findings to suggest that the definition of a health system resilient to infectious disease shocks is a well-prepared system capable of anticipating and responding to imminent infectious disease emergencies; able to withstand such emergencies without severe adverse effects; capable of adapting and maintaining the provision of essential services while enhancing emergency response; and capable of recovering promptly after the emergency to continue delivering critical services. This description highlights four essential capacities that must be actively maintained to define a resilient health system, including preparedness, absorptive capacity, adaptive capacity, and recovery.

Previous definitions of health system resilience reference three capacities: absorptive, adaptive, and transformative (104). The definition outlined in our study encompasses preparedness and recovery; however, it does not include the concept of transformation capacities. Studies have emphasized the significance of preparedness as an essential component of resilience (105).

Another article observed that the destruction of health systems caused by COVID-19 was attributable to the inadequate preparation of many systems, characterized by a deficiency in robust surveillance and testing capabilities (106). Preparedness activities, such as establishing systems, stockpiling medical countermeasures and other essential supplies, and conducting capacity-building initiatives, are crucial for enhancing the resilience of the healthcare system. Preparedness has been linked to the absorptive capacity, as well as the adaptive capacities of a health system (107). However, we argue that, to the extent that the benefit of preparedness includes prevention efforts—beyond just absorbing a shock—it should be considered a separate component capacity.

Our definition references recovery, which denotes returning to the pre-crisis state (108). This requires a reconsideration towards developing transformative capacities, which is a function of learning from previous crises and constructing a system capable of better responding to future shocks (109,110). According to our study, the absorptive capacity of a health system refers to its ability to withstand a crisis, develop effective coping strategies, and maintain uninterrupted services. This aligns with the definitions found in several articles (105,107).

The attributes of resilient systems include leadership and governance, community engagement, health service delivery, health education and promotion, surveillance and laboratory, innovations, and continuous learning and adaptation. Earlier studies identified five attributes of a resilient health system: aware, integrated, diverse, self-regulating, and adaptive (16). According to the article, these attributes, also called dimensions, are defined as: aware — ability to detect risks early and understand its own strengths and vulnerabilities; diverse — offers a broad range of services and delivery models to meet health needs; self-regulating — the system can isolate threats and maintain core functions during crises; integrated — coordinates across sectors and engages communities to build trust and accountability; and adaptive — the system can learn from crises and adjust as may be required. There are considerable overlaps in the attributes between the subjects of our study and those discussed in the article; however, we will elaborate on community engagement and health education and promotion, which do not appear to be sufficiently addressed in the five dimensions.

The dimension “integrated” relates to the engagement of community leaders; however, numerous countries in Africa have health system frameworks that are embedded within the community, necessitating not merely involvement but active participation (111). Community healthcare workers engage in the provision of primary healthcare services and serve as more than just intermediaries between the health system and the community (100). Recognizing the role of the community as a major stakeholder in the development of resilient health systems, there have been calls for their inclusion as a fundamental component of the health system—beyond the traditional six building blocks outlined by the World Health Organization (112). In the specific instances of West Africa and Uganda, community healthcare workers fulfilled essential roles in managing outbreaks and in the early detection of threats to facilitate an effective response (75,96). Regarding community engagement, health education and promotion empower community members to take responsibility for their own health. Community healthcare workers and a community empowered with information on health are essential to the achievement of Universal Health Coverage, which resilient health systems strive to attain (113).

The World Health Organization associates the resilience of health systems with health security and universal health coverage; therefore, initiatives to strengthen resilient health systems contribute to a more secure global health environment, ensuring that healthcare is accessible to all at all times (5). The World Health Organization’s call for nations to develop resilient health systems is therefore one that must be addressed with urgency (114). Heeding the call to build resilient health systems necessitates a refined understanding of the concept. Resilience is inherently a dynamic notion that must consider the experiences of individual health systems; therefore, its implementation should not be prescriptive but shaped by the specific context (115).

This review exclusively considered articles published after 1980, using an arbitrary cutoff, acknowledging that valuable studies on health system resilience may have been conducted prior to this date. Additionally, we recognize that during the screening of titles and abstracts, some articles that could have been included might have been unintentionally excluded.

## Conclusion

The description of health system resilience to infectious diseases emphasizes preparedness, absorptive and adaptive capacities, as well as recovery. Our study underscores seven attributes, five of which are thoroughly represented by the dimensions of a resilient health system: awareness, integration, diversity, adaptability, and self-regulation. Nevertheless, it is our considered view that our research introduces community engagement and health education and promotion as essential attributes of a resilient health system. The implications of these findings is that preparedness needs to be integrated in the definition of health system resilience, highlighting the role of prevention in the concept of resilience. Considering the operational definition of recovery that encompasses bouncing back to pre-shock levels, policymakers should explore the incorporation of lessons learned to strengthen health systems beyond their pre-outbreak state. We recommend these elements of resilience from these findings be included in the frameworks of health system resilience within the continent. Very importantly, these findings provide the foundation for the development of tools for the measurement and monitoring of health system resilience. Future research can build on the findings of this review to refine the definition and theories of health system resilience.

## Data Availability

All the data are submitted in the Supporting Information attached

## Author contributions

DO conceptualized the study, developed the search strategy, conducted the search in databases, extracted data, analyzed the data, and drafted the manuscript; MJ led the development of the search strategy and searching the databases; SA was involved in both title-abstract and full-text screening; MV, RM, ER, VN, FO, UNM, ARA, FF and SNK provided technical guidance in the development of the study, and all the processes including drafting the manuscript; CLO, EBN, SN and EA were involved in data analysis, review of the manuscript and editing.

## Acknowledgments

The authors extend their gratitude to the Norwegian Institute of Public Health and the Uganda National Institute of Public Health for their continued support towards the development of this review. VN was supported by an EDCTP2 Senior Fellowship (Grant No.: TMA2018SF-2479).

## Supporting documents

S1 Checklist: PRISMA Reporting Guidelines

S1 Text: Search strategy and search log

S1 Table: Excluded records

S2 Table: Characteristics of included records

S3 Table: Quality appraisal of qualitative studies using CASP

S4 Table: Quality appraisal of mixed-methods studies using MMAT

